# Systematic benchmarking demonstrates large language models have not reached the diagnostic accuracy of traditional rare-disease decision support tools

**DOI:** 10.1101/2024.07.22.24310816

**Authors:** Justin T Reese, Leonardo Chimirri, Yasemin Bridges, Daniel Danis, J Harry Caufield, Michael A. Gargano, Carlo Kroll, Andrew Schmeder, Fengchen Liu, Kyran Wissink, Julie A McMurry, Adam SL Graefe, Enock Niyonkuru, Daniel R Korn, Elena Casiraghi, Giorgio Valentini, Julius OB Jacobsen, Melissa Haendel, Damian Smedley, Christopher J Mungall, Peter N Robinson

## Abstract

Large language models (LLMs) show promise in supporting differential diagnosis, but their performance is challenging to evaluate due to the unstructured nature of their responses and their accuracy compared to existing diagnostic tools is not well characterized. To assess the current capabilities of LLMs to diagnose genetic diseases, we benchmarked these models on 5,213 case reports using the Phenopacket Schema, the Human Phenotype Ontology and Mondo disease ontology. Prompts generated from each phenopacket were sent to seven LLMs, including four generalist models and three LLMs specialized for medical applications. The same phenopackets were used as input to a widely used diagnostic tool, Exomiser, in phenotype-only mode. The best LLM ranked the correct diagnosis first in 23.6% of cases, whereas Exomiser did so in 35.5% of cases. While the performance of LLMs for supporting differential diagnosis has been improving, it has not reached the level of commonly used traditional bioinformatics tools. Future research is needed to determine the best approach to incorporate LLMs into diagnostic pipelines.

## Introduction

Rare diseases, collectively, are not rare: over 10,000 rare diseases (RDs) have been identified to date, together affecting between 3.5% and 8% of the population. Furthermore, affected individuals often experience a diagnostic odyssey lasting 5-7 years.^1,2^ Computational comparison of the phenotypic abnormalities of individuals with the phenotypic profiles of disease is a cornerstone of the diagnostic workup of individuals with suspected RD. Numerous algorithms leverage descriptions of clinical manifestations formulated as Human Phenotype Ontology (HPO) terms to search for diseases with similar manifestations.^3–7^ Recently, the performance of large language models (LLMs) in supporting the differential diagnostic process has been assessed by many groups. LLMs are general-purpose artificial intelligence models that can be applied to numerous tasks across diverse domains. LLMs have been shown to be useful for many clinical tasks, such as charting, medication review, or clinical trial matching.^8,9^ For differential diagnostic support, LLMs can be prompted to return a ranked list of potential diagnoses based on narrative text describing a patient’s features.^10^

We identified 36 previous publications that evaluated the performance of LLMs on differential diagnostic challenges using text prompts (Supplemental Table 1). Thirty-four of 36 publications we identified involved human curation, usually by physicians, to compare the response of LLMs (most frequently, OpenAI’s GPT models) to the correct diagnosis recorded in the original source (Figure 1 and Supplemental Table S1). These studies analyzed cohorts of between 6 and 9681 cases (median 78), often from published vignettes intended for medical education, such as the *Case Studies* of New England Journal of Medicine (NEJM),^10–13^ the *Diagnosis Please* quizzes from the journal Radiology,^14^ and JAMA Ophthalmology *Clinical Challenges*.^15^ The reported performance varied widely, even for studies using the same input data such as the NEJM Case Studies (Figure 1). We reasoned that the variability could be partially due to subjective judgements as to whether an LLM response exactly matched the correct diagnosis.

**Figure 1.**
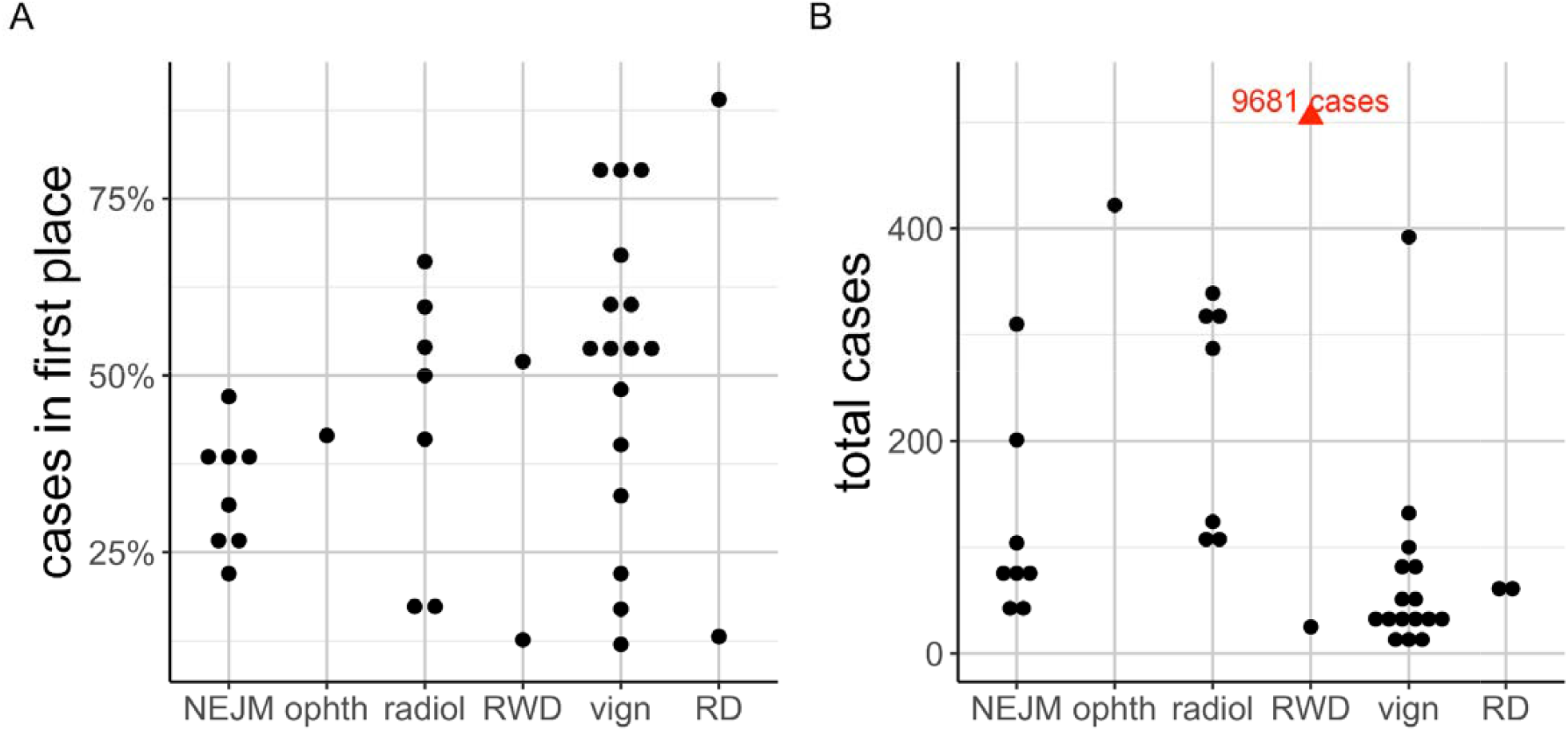
Accuracy of LLMs in differential diagnostic challenges. Summary of performance in 33 previously published studies that reported the percentage of cases in which the correct diagnosis was placed at rank 1 by the LLM. Cohorts were derived from multiple sources including published clinical vignettes (vign), New England Journal of Medicine case reports or quizzes (NEJM), JAMA Ophthalmology Clinical Challenges (ophth), and case reports including clinical data and radiology reports in text form (radiol), and one cohort of real-world data (RWD; 6 patients), and rare disease (RD). Details are available in Supplemental Table 1.

To provide an estimate of the performance of LLMs for differential diagnostics in human genetics, we assembled data from 5213 individuals with a previously solved diagnosis; each case was structured as a phenopacket,^22^ a Global Alliance for Genomics and Health (GA4GH) and ISO standard. Each phenopacket provides a structured representation of Human Phenotype Ontology (HPO) terms representing the signs, symptoms, abnormal imaging findings and laboratory test results that were observed or excluded in a single patient. Collectively, the diagnoses spanned 378 unique Mendelian or chromosomal diseases; the phenopacket encodes these as (OMIM)^23^ identifiers. The identified causal variants, while part of the phenopacket, were not used in the current analysis. We programmatically generated prompts from the phenopackets using a standard template (all phenopackets and prompts are available via Zenodo - see data availability section). We developed an approach to assessment that mitigates some common challenges in fairly assessing LLM performance and comparing it to that of standard bioinformatic tools. We found that Exomiser clearly outperforms the current generation of LLM tools.

## Methods

### Study design and data

We evaluated the performance of LLMs in differential diagnosis using 5213 computational case reports formatted as GA4GH phenopackets taken from the phenopacket-store repository (version 0.14).^24^ The case reports describe 378 Mendelian and chromosomal diseases associated with 336 genes. Each phenopacket contains information derived from published case or cohort reports from a total of 726 different publications. The diagnosis indicated in the original publication was recorded but not used in generating the prompt. A total of 2975 distinct HPO terms were used, with an average of 16 HPO terms per case.

This study was deemed exempt from local IRB approval as it did not meet the criteria for human subjects research, because the cases were curated from publicly available articles.

### Retrieval of relevant literature

A search was conducted in PubMed to retrieve articles that describe the use of LLMs for differential diagnostics. The search string was:

> (“GPT”[Title/Abstract] OR “LLM”[Title/Abstract] OR “Large language model”[Title/Abstract])
>
> AND
>
> (“differential diagnostics”[Title/Abstract] OR “differential diagnosis”[Title/Abstract])
>
> AND
>
> (“2023”[Date - Create] : “3000”[Date - Create])

This search was performed on Oct 18, 2024, and returned 63 articles.

This list was further refined by selecting only articles that described the application of one or more LLMs to perform differential diagnostic analysis on a cohort of clinical cases. Further, for better comparability, we only retained publications in which the rate of placing the correct diagnosis in rank 1 was reported. For instance, we omitted one publication because only the rate of the correct diagnosis in the top three candidates was reported.^25^ The reference lists of chosen publications were scanned to identify additional articles. A full list of included articles is provided in Supplemental Table 1.

### Computational generation of prompts for LLMs

We constructed software, *phenopacket2prompt*, to convert case data in GA4GH phenopacket format to prompts suitable for use with LLMs to generate differential diagnoses. By parsing phenopackets, the software extracts relevant data such as age, gender, phenotypic features that were observed and excluded in the patient, and onset information. This information is then used together with a programmatic template to generate a clinical narrative suitable for LLMs. The template first specifies the sex of the individual, the age of onset, and the age at last examination, and then lists the HPO terms that represent observed or excluded clinical features. If available, separate lists are included for different ages of examination. The software is implemented as a command-line Java application and is freely available on GitHub under an open source MIT license at https://github.com/monarch-initiative/phenopacket2prompt. For some models (Meditron-70B, Meditron3-70B, Medfound-176B, Gemini Flash 2.0), we assessed prompts from phenopacket2prompt and also bespoke prompts that conform to those described previously for each model(Liu et al. 2025; Chen et al. 2023). Results reported for these models here are for the better-performing bespoke prompts. Example prompts (Supplemental Tables S2-8) and the full set of prompts and responses for all models https://zenodo.org/records/15324355) are provided.

### Generating differential diagnoses

#### Exomiser

Exomiser was used to generate differential diagnoses as follows. Exomiser version 14.0.1 was downloaded from https://github.com/exomiser/Exomiser/releases/download/14.0.1/exomiser-cli-14.0.1-distribution.zip, and the 2406 version of the Exomiser data release was downloaded from: https://data.monarchinitiative.org/exomiser/data/2406_phenotype.zip https://data.monarchinitiative.org/exomiser/data/2406_hg19.zip

Exomiser can be run with exome or genome sequence data and HPO terms as input, or with only HPO terms (phenotype only mody). Exomiser was applied in phenotype only mode to generate a differential diagnosis (comprising ranked lists of OMIM or Orphanet IDs) for each phenopacket.

#### LLMs

The prompts generated as described above were provided to o1-preview (version o1-preview-2024-09-12), o1 mini (version o1-mini-2024-09-12), and GPT-4o (version gpt-4o-2024-08-06), Gemini Flash 2.0 (gemini-2.0-flash-001), Meditron-70B (version 2023-12-07), Meditron3-70B (version 2024-07-11), and Medfound-176B (version 2024-11-044) to generate differential diagnoses. For o1-preview, o1 mini, and GPT-4o, prompts were sent to the OpenAI API (https://platform.openai.com/docs/api-reference), and for Gemini Flash 2.0 prompts were sent to the Gemini API (https://ai.google.dev/gemini-api/docs). Meditron-70B, Meditron3-70B, and Medfound-176B models were downloaded from Huggingface (https://huggingface.co/epfl-llm/meditron-70b, https://huggingface.co/OpenMeditron/Meditron3-70B, https://huggingface.co/medicalai/MedFound-176B).

Meditron and Medfound models were deployed on an HPC cluster using a compute node equipped with four H100 GPUs. We leveraged the inferencing code provided by Meditron (https://github.com/epfLLM/meditron) to both configure default model hyperparameters and scale inferencing using the vLLM (https://github.com/vllm-project/vllm) inference engine.

For each case, each item in the differential diagnosis generated by the LLM was converted to Mondo Disease Ontology terms as follows: first, items that matched exactly to a Mondo disease label or synonym were assigned Mondo terms using the Ontology Access Kit (OAK, https://github.com/INCATools/ontology-access-kit), and the remaining items were assigned the best matching Mondo term above an empirically determined threshold using CurateGPT (https://github.com/monarch-initiative/curategpt). 94.0% and 95.6% of items in the differential diagnoses were assigned Mondo terms.

### Scoring differential diagnoses

For each case, each item in the differential diagnosis was scored as correct or incorrect using OAK as follows. An item from the differential diagnosis was considered correct if the disease identifier for the diagnosis matched the identifier for gold standard diagnosis from the case report exactly, or was mapped as equivalent to the identifier of the gold standard diagnosis in Mondo, or if the gold standard diagnosis was a close descendant of the LLM’s diagnosis identifier in Mondo (Figure 2). Thus, more general items such as *geleophysic dysplasia* (MONDO:0000127) are considered as correct diagnoses for more specific items that are subsumed by the general one, such as *geleophysic dysplasia 2* (MONDO:0013612). For each case, the rank of the correct diagnosis, if present, was recorded.

**Figure 2.**
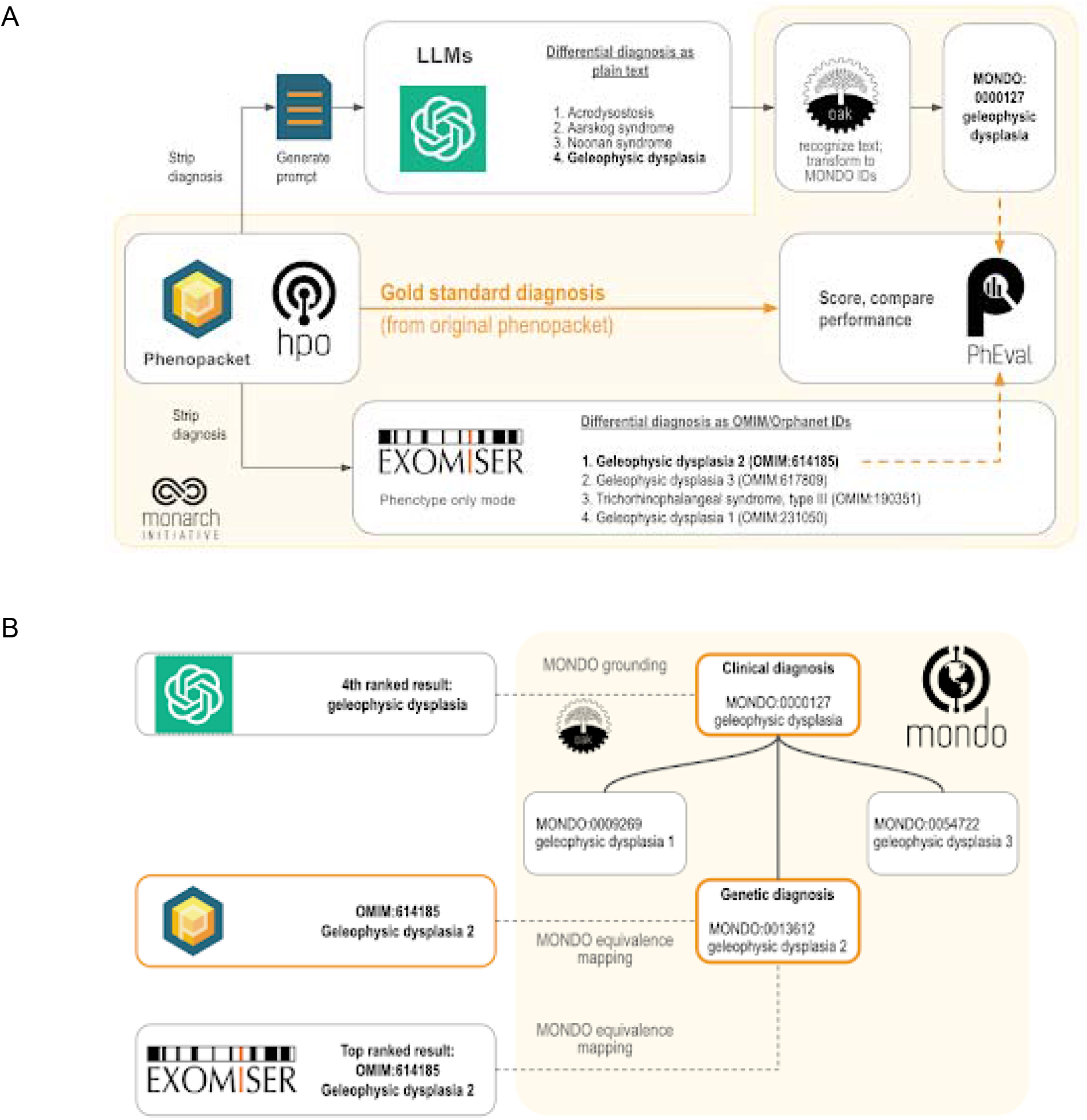
Workflow for comparing the accuracy of differential diagnoses from Exomiser and LLMs. Each phenopacket in the cohort of 5213 cases is used to generate a differential diagnosis with LLMs or Exomiser. **(A)** The phenopacket is used to generate a prompt containing case data (age, gender, observed and excluded phenotypic features, and the onset of phenotypic features, if present). Each item in the LLM response is converted to Mondo disease identifiers using concept recognition software (OAK). Exomiser uses the same phenopacket to generate a differential diagnosis comprising OMIM or Orphanet disease identifiers from the Exomiser dataset. The rank of the correct diagnosis, if present, is determined by comparing them to the gold standard diagnosis from the phenopacket using an ontology-based strategy. **(B)** Ontology-based strategy for identifying correct diagnoses (orange) using Mondo. Items in the differential diagnoses from LLM (typically clinical diagnoses, e.g. geleophysic dysplasia) are grounded to Mondo, and items from Exomiser (typically genetic diagnoses, e.g. geleophysic dysplasia 2) and the correct diagnosis from the phenopacket are aligned to Mondo using equivalence mappings in Mondo. An item in a differential diagnosis is considered correct if it matches the diagnosis from the phenopacket (OMIM:614185 geleophysic dysplasia 2), matches an equivalent Mondo disease (MONDO:0013612 geleophysic dysplasia 2), or matches a more general Mondo disease with a descendant that is a correct diagnosis (MONDO:0000127 geleophysic dysplasia).

## Results

In this work, we develop an evaluation strategy to estimate the expected performance of LLMs as compared to Exomiser, a specialist software tool for differential diagnostic support in human genetics. It is necessary to find a common approach to measuring the accuracy of responses of both tools - LLMs respond in free text, whereas Exomiser provides a ranked list of diagnoses encoded as Online Mendelian Inheritance in Man (OMIM) or Orphanet codes (Orphacodes). Additionally, many tools in human genetics, including Exomiser, provide precise diagnoses, (e.g., Bardet-Biedl syndrome-14, OMIM:615991). Since LLMs are not currently designed to prioritize genetic findings, we performed our comparison using only phenotypic findings. We therefore developed an evaluation strategy that regards groups of diseases that are clinically identical or closely related as equivalent (in this example, all genetic forms of Bardet-Biedl syndrome would be regarded as equivalent for the purposes of ranking the correct answer).

In the following, we present our analysis of current challenges in fairly assessing the performance of LLMs in differential diagnostic tasks, describe our approach to assessing LLM performance in rare-disease diagnostic support, and finally present our analysis of seven LLMs as compared to Exomiser over a total of 5,213 cases.

### Evaluation strategy

Although conceptually simple, evaluation of the differential diagnosis returned by an LLM requires that the curator have specialized medical knowledge of the disease in question. For instance, in case Case 2-2021 of the NEJM Case Record series that has been used by multiple groups to assess LLM performance,^10^ the final diagnosis was given as “pregnancy-associated myocardial infarction, probably due to spontaneous coronary-artery dissection”.^16^ In our analysis of this case,^17^ GPT-4 returned answers including “peripartum cardiomyopathy” and “heart failure secondary to severe pre-eclampsia,” both of which are cardiovascular complications following delivery, but neither of which is correct. As a second example, in case 16-2021, the final diagnosis was “*Staphylococcus aureus* bacteremia and infection of a vascular graft”.^18^ In our analysis of this case, the diagnosis at rank 4, “infective endocarditis affecting the aortic valve and causing referred abdominal pain,” was similar to the correct diagnosis but does not identify the causative agent (*i*.*e*., *S. aureus*). In a study on the NEJM cases, two scorers agreed on only 66% of scores in 80 NEJM cases.^10^ Therefore, the practical utility of LLM analysis may be limited by the varying ability of human users to interpret the responses, and manual curation that may influence the measured performance of LLMs. For this reason, we concluded that a computational approach to evaluate the responses of LLMs such as GPT-4 by assigning them to specific disease entities (Mondo ontology terms) would provide a more realistic assessment of the performance of these models in diagnostic applications.

Two recent LLM studies by Kim et al.^19^ and by Flaharty et al.^20^ focus on diagnosing rare genetic disease (RD). The Kim et al. study requested the LLMs to return gene symbols rather than disease names or codes,^19^ which obviates the need to manually check for equivalence of potentially synonymous disease names; however, LLMs are known to frequently return erroneous gene and ontology identifiers (a form of hallucination).^21^ This study of the performance of several LLMs on 276 published case reports showed that GPT-4 had the best performance, placing the correct gene symbol (used as a proxy for the diagnosis) of 13.9% of cases within the top ten ranked diagnoses.

However, there are several potential shortcomings with this approach. Predicting disease genes is arguably more challenging than predicting diseases, because one gene may be associated with many diseases, and the same (clinically defined) disease may be associated with many genes. For example, the authors included genes like *LMNA*, which is linked to 11 distinct diseases in Online Mendelian Inheritance in Man (OMIM), and *IFT172*, which is associated with three diseases, of which one (Bardet-Biedl syndrome) is a genetically complex condition that can result from pathogenic variants in any of at least 22 different genes. In this study, the correct gene was placed in the top ten ranked candidates in 11.7% of cases; the rank-1 performance was not reported.

In contrast, the study by Flaharty et al. investigated 63 genetic conditions and compared the performance of prompts composed with medical and lay language.^20^ The 63 cases were created with 2-5 characteristic phenotype terms each and were described by the authors as “textbook-like descriptions.” This work involved a similar manual assessment step to determine if the response of the LLM was correct; for instance, in one case, the authors assigned “Riley-Day syndrome’’ to its synonym ‘‘hereditary sensory and autonomic neuropathy, type III’’, and reported that some LLM responses required additional discussion among the graders. Among the LLMs to date, GPT models have been shown to have the best general performance in differential diagnosis (Figure 1; see references in Supplemental Table 1).

### An approach for assessing LLM and Exomiser Performance in Differential Diagnostic Support

We analyzed the same 5213 cases to compare performance between seven different LLMs and a traditional bioinformatics tool, Exomiser, which was developed by the Monarch Initiative. Exomiser was shown to be the best performing diagnostic tool on 100,000 Genomes Project data^27^ and is widely used in diagnostic labs, large-scale disease sequencing projects and national healthcare services such as the UK’s Genomic Medicine Service. For the comparison herein, we used Exomiser in ‘phenotype only’ mode. To mitigate the potential of bias implied by manual comparison of LLM responses to the expected correct result, we developed an approach to programmatically map (i.e. ‘ground’) responses of the LLM to terms from the Monarch Initiative’s Mondo disease ontology (https://github.com/monarch-initiative/mondo), which provides a comprehensive and standardized framework used for the classification of human diseases that integrates various disease classification systems, and thereby provides a unified approach to disease nomenclature.^28^ For genetically heterogeneous diseases such as geleophysic dysplasia, we used Mondo to “roll up” groups of diseases (e.g., MONDO:0000127 geleophysic dysplasia subtypes 1 through 3 of geleophysic dysplasia).

A total of seven different LLMs were investigated (Table 1). We presented the LLM with these prompts generated from phenopackets (Figure 2; Supplemental Tables S2-S8). We prompted the model to return a differential diagnosis as a list of disease names and determined the rank of the correct diagnosis in these lists, if present. Our strategy employs the Mondo ontology to count clinical diagnoses (e.g., geleophysic dysplasia) as correct, in addition to the the original precise genetic diagnosis (e.g., geleophysic dysplasia 2) since no genetic information was provided to any diagnostic tool in this experiment. We used the PhEval evaluation framework^29^ to rigorously benchmark results.

**Table 1.** Summary of LLMs evaluated for diagnostic accuracy in this study.

| LLM | Description |
| --- | --- |
| GPT o1 preview | Larger reasoning model from OpenAI<sup>30</sup> |
| GPT o1 mini | Smaller reasoning model from OpenAI<sup>30</sup> |
| GPT-4o | Non-reasoning model from OpenAI<sup>31</sup> |
| Gemini 2.0 Flash | Non-reasoning model from Google<sup>32</sup> |
| Meditron-70B | Medical foundation model from EPFL<sup>33</sup> |
| Meditron3-70B | Updated version of Meditron-70B from EPFL<sup>33</sup> |
| Medfound-176B | Medical foundation model<sup>26</sup> |

### Evaluation on 5213 cases

The diagnostic accuracy of Exomiser was greater than any of the LLMs tested. Exomiser placed the correct diagnosis in the first rank in 35.5% of cases, compared with 23.6% for the best performing LLM (o1 preview). Similarly, Exomiser placed the correct diagnosis in the top 3 ranks of the differential diagnosis in 46.3% of cases and in the top 10 ranks in 58.5% of cases, compared with 31.2% in the top 3 ranks and 36.8% in the top 10 ranks for the best performing LLM (o1 preview). Among the medical foundation models, Meditron3-70B performed best, placing the correct diagnosis in the top 1, 3 and 10 ranks 19%, 28%, and 31%, respectively, but its accuracy was lower than all general foundation models we tested. Medfound-176B performed the worst, placing the correct diagnosis in the top 1,3, and 10 ranks for only 0.1%, 0.23%, and 0.25% of cases (Figure 3).

**Figure 3.**
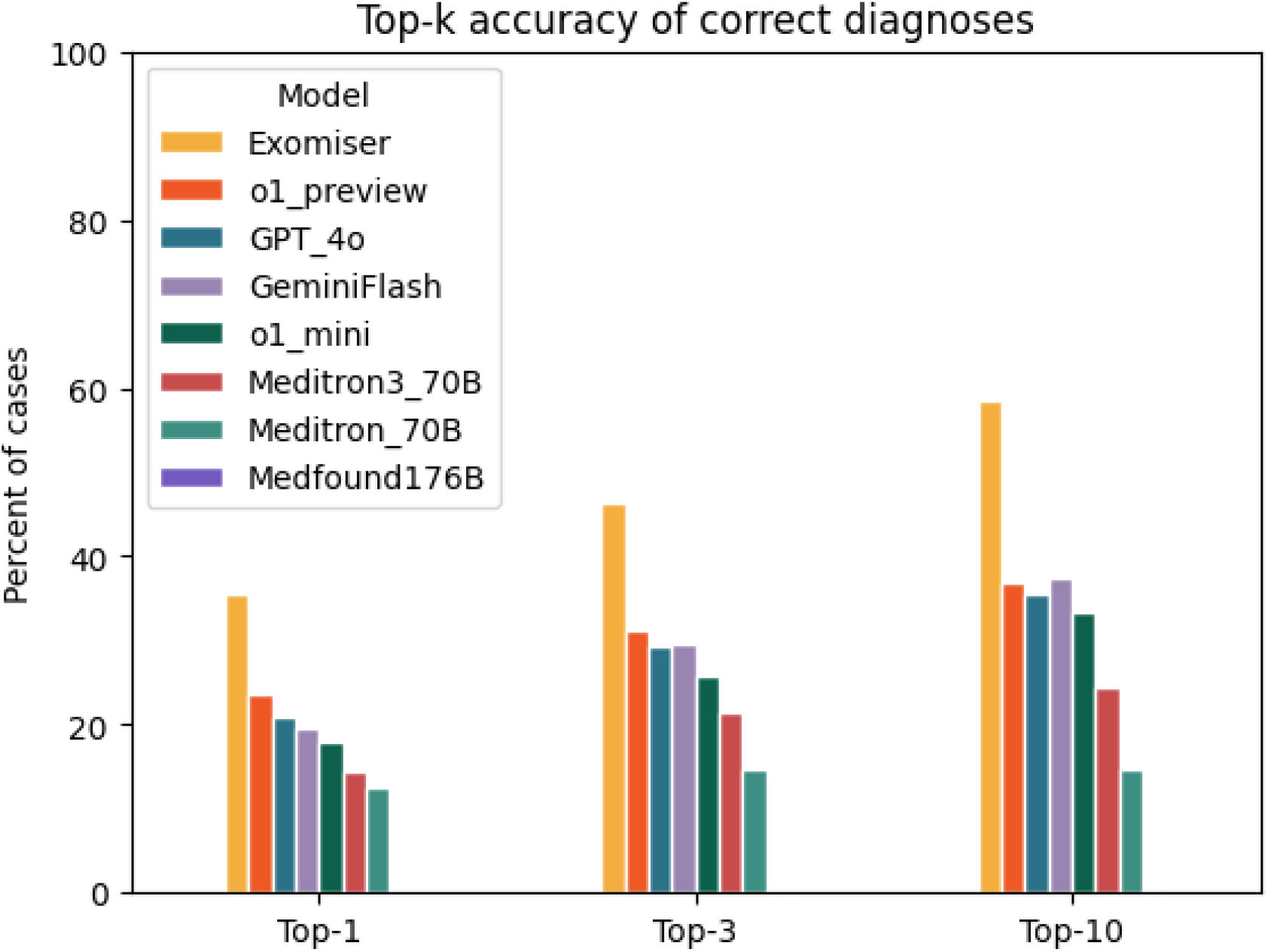
Accuracy of Exomiser compared with large language models in differential diagnostic challenges. The bar chart shows the percentage of cases of the current cohort of 5213 cases in which Exomiser, o1 preview, GPT-4o, Gemini Flash, o1 mini, Meditron3-70B, Meditron-70B, and Medfou d-176B returned a correct diagnosis at rank 1 (Top-1), within the top 3 (Top-3), or within the top 10 ranks (Top-10). Medfound-176B displayed a performance of 0.25% or less in all tests and is not visible in the bar chart.

## Discussion

Previous studies that evaluated the performance of LLMs in the area of decision support for differential diagnosis have had relatively small sample sizes and have employed manual and subjective evaluation of whether LLM responses match the correct diagnosis. Our analysis uses an ontology-based strategy that eliminates subjective choices as to whether an unstructured response returned by an LLM contains the correct diagnosis, and provides a realistic estimate of the expected performance of LLMs over a broad range of rare diseases. In our analysis, the best performing LLM (o1 preview) was able to place the correct diagnosis in the first ten ranks in 36.8% of the 5213 cases, much higher than in the study using gene symbols (12.11%); the performance was worse than for the 63 textbook-like disease descriptions (81%–89% top-10 accuracy). In our study, the performance of Exomiser was substantially better than that of the best LLM using only phenotype data. Additionally, Exomiser is designed to work with both phenotypic and whole-exome or whole-genome data; on 4877 molecularly diagnosed cases from the 100,000 Genome Project, Exomiser prioritized the correct gene in the top, top 3, and top 10 ranked candidates 82.6%, 91.3%, and 93.6% of the time.^34^ We did not test the utility of providing the LLM with genomic information in the current analysis; including genetic information would require additional measures to ensure privacy if used for actual patients.

Limitations of our study include the fact that the representation of the clinical phenotypes with HPO terms in the phenopackets may have been incomplete or inaccurate. Additionally, the description of the clinical features in the publications from which the phenopackets were derived may have been incomplete. We did not undertake fine-tuning or prompt-tuning in this analysis; these procedures may increase performance on specific clinical decision-making tasks.^35^ A recent study that employs a likelihood ratio method combined with retrieval augmented prompt generation, followed by querying of GPT-3.5-turbo, showed a top-ten performance of 69.33% (165 out of 238 cases) for the true diagnoses in the top 10; however, the same cases were used to train the model, so that the expected performance on new cases and diseases remains to be characterized.^21^ While medical foundation models (Meditron3-70B, Meditron-70B, and Medfound-176B) all performed more poorly than general-purpose LLMs, it is possible that fine-tuning on diagnostic tasks may improve performance. It is possible that the larger model sizes and resources used to train the general-purpose LLMs are related to the differential performance. A version of Medfound-176B that has been fine-tuned for diagnostic use has been described previously,^26^ but is currently not available for benchmarking.

In conclusion, we present the largest reported study on the differential diagnostic capabilities of the GPT family of LLMs for rare genetic disease, the LLM that is the current best in class for a variety of medical applications. Our analysis approach was designed to minimize variability and subjective choices in evaluation, and thereby provides a realistic estimate of the performance of GPT in rare-disease differential diagnostics, and shows that the performance of GPT for RD differential diagnostic support is currently clearly inferior to that of a commonly used traditional bioinformatics tool, Exomiser. Future work will be required to determine how to design and integrate LLMs into diagnostic pipelines for RD genomic diagnostics.

## Data Availability

All data produced are available online on Zenodo at: https://zenodo.org/records/15324355.

## Acknowledgements

We acknowledge the role of the GA4GH in providing the framework in which the Phenopacket Schema was developed from 2016 to 2022. This work was funded by grants 5R01HD103805-03 from the National Institute of Child Health and Human Development, 5R24OD011883-06 from the Office of the Director of the National Institutes of Health (NIH), and 5RM1 HG010860-03 from the National Human Genome Research Institute. Additional support was provided by the Alexander von Humboldt Foundation, the Director, Office of Science, Office of Basic Energy Sciences of the U.S. Department of Energy Contract No. DE-AC02-05CH11231, and FAIR (Future Artificial Intelligence Research) project, funded by the NextGenerationEU program within the PNRR-PE-AI scheme (M4C2, Investment 1.3, Line on Artificial Intelligence) -AIDH – FAIR - PE0000013.

## Data and code availability

### Data availability

The GA4GH phenopacket case data (derived from published case reports), prompts generated by phenopacket2prompt, and differential diagnoses generated by LLMs and Exomiser are available on Zenodo: https://zenodo.org/records/15324355. All data collected for the study is available to all under an open source CC-BY License.

### Code availability

phenopacket2prompt is freely available on GitHub under an open source MIT license at https://github.com/monarch-initiative/phenopacket2prompt. The grounding and evaluation software is freely available on GitHub under an open-source BSD 3-Clause License at https://github.com/monarch-initiative/pheval.llm. OAK is freely available on GitHub under an open-source Apache 2 license.

### Reporting

Reporting in this study followed Consolidated Reporting Guidelines for Prognostic and Diagnostic Machine Learning Modeling Studies.^36^

## References

1. Vandeborne, L., van Overbeeke, E., Dooms, M., De Beleyr, B., and Huys, I. (2019). Information needs of physicians regarding the diagnosis of rare diseases: a questionnaire-based study in Belgium. Orphanet J. Rare Dis. 14, 99.

2. Haendel, M., Vasilevsky, N., Unni, D., Bologa, C., Harris, N., Rehm, H., Hamosh, A., Baynam, G., Groza, T., McMurry, J., et al. (2020). How many rare diseases are there? Nat. Rev. Drug Discov. 19, 77–78.

3. Smedley, D., and Robinson, P.N. (2015). Phenotype-driven strategies for exome prioritization of human Mendelian disease genes. Genome Med. 7, 81.

4. Köhler, S., Schulz, M.H., Krawitz, P., Bauer, S., Dölken, S., Ott, C.E., Mundlos, C., Horn, D., Mundlos, S., and Robinson, P.N. (2009). Clinical diagnostics in human genetics with semantic similarity searches in ontologies. Am. J. Hum. Genet. 85, 457–464.

5. Bauer, S., Köhler, S., Schulz, M.H., and Robinson, P.N. (2012). Bayesian ontology querying for accurate and noise-tolerant semantic searches. Bioinformatics 28, 2502–2508.

6. Yuan, X., Wang, J., Dai, B., Sun, Y., Zhang, K., Chen, F., Peng, Q., Huang, Y., Zhang, X., Chen, J., et al. (2022). Evaluation of phenotype-driven gene prioritization methods for Mendelian diseases. Brief. Bioinform. 23,.

7. Beckwith, M.A., Danis, D., Bridges, Y., Jacobsen, J.O.B., Smedley, D., and Robinson, P.N. (2025). Leveraging clinical intuition to improve accuracy of phenotype-driven prioritization. Genet. Med. 27, 101292.

8. Singhal, K., Azizi, S., Tu, T., Mahdavi, S.S., Wei, J., Chung, H.W., Scales, N., Tanwani, A., Cole-Lewis, H., Pfohl, S., et al. (2023). Large language models encode clinical knowledge. Nature 620, 172–180.

9. Moor, M., Banerjee, O., Abad, Z.S.H., Krumholz, H.M., Leskovec, J., Topol, E.J., and Rajpurkar, P. (2023). Foundation models for generalist medical artificial intelligence. Nature 616, 259–265.

10. Kanjee, Z., Crowe, B., and Rodman, A. (2023). Accuracy of a Generative Artificial Intelligence Model in a Complex Diagnostic Challenge. JAMA.

11. Abdullahi, T., Singh, R., and Eickhoff, C. (2024). Learning to Make Rare and Complex Diagnoses With Generative AI Assistance: Qualitative Study of Popular Large Language Models. JMIR Med Educ 10, e51391.

12. Ríos-Hoyo, A., Shan, N.L., Li, A., Pearson, A.T., Pusztai, L., and Howard, F.M. (2024). Evaluation of large language models as a diagnostic aid for complex medical cases. Front. Med. 11, 1380148.

13. Chiu, W.H.K., Ko, W.S.K., Cho, W.C.S., Hui, S.Y.J., Chan, W.C.L., and Kuo, M.D. (2024). Evaluating the Diagnostic Performance of Large Language Models on Complex Multimodal Medical Cases. J. Med. Internet Res. 26, e53724.

14. Ueda, D., Mitsuyama, Y., Takita, H., Horiuchi, D., Walston, S.L., Tatekawa, H., and Miki, Y. (2023). ChatGPT’s Diagnostic Performance from Patient History and Imaging Findings on the Diagnosis Please Quizzes. Radiology 308, e231040.

15. Milad, D., Antaki, F., Milad, J., Farah, A., Khairy, T., Mikhail, D., Giguère, C.-É., Touma, S., Bernstein, A., Szigiato, A.-A., et al. (2024). Assessing the medical reasoning skills of GPT-4 in complex ophthalmology cases. Br. J. Ophthalmol.

16. Scott, N.S., Thomas, S.S., DeFaria Yeh, D., Fox, A.S., and Smith, R.N. (2021). Case 2-2021: A 26-Year-Old Pregnant Woman with Ventricular Tachycardia and Shock. N. Engl. J. Med. 384, 272– 282.

17. Reese, J.T., Danis, D., Caufield, J.H., Groza, T., Casiraghi, E., Valentini, G., Mungall, C.J., and Robinson, P.N. (2024). On the limitations of large language models in clinical diagnosis. medRxiv.

18. Dua, A., Sutphin, P.D., Siedner, M.J., and Moran, J. (2021). Case 16-2021: A 37-Year-Old Woman with Abdominal Pain and Aortic Dilatation. N. Engl. J. Med. 384, 2054–2063.

19. Kim, J., Wang, K., Weng, C., and Liu, C. (2024). Assessing the utility of large language models for phenotype-driven gene prioritization in the diagnosis of rare genetic disease. Am. J. Hum. Genet. 111, 2190–2202.

20. Flaharty, K.A., Hu, P., Hanchard, S.L., Ripper, M.E., Duong, D., Waikel, R.L., and Solomon, B.D. (2024). Evaluating large language models on medical, lay-language, and self-reported descriptions of genetic conditions. Am. J. Hum. Genet. 111, 1819–1833.

21. Yang, J., Shu, L., Duan, H., and Li, H. (2024). RDguru: A conversational intelligent agent for rare diseases. IEEE J. Biomed. Health Inform. PP,.

22. Jacobsen, J.O.B., Baudis, M., Baynam, G.S., Beckmann, J.S., Beltran, S., Buske, O.J., Callahan, T.J., Chute, C.G., Courtot, M., Danis, D., et al. (2022). The GA4GH Phenopacket schema defines a computable representation of clinical data. Nat. Biotechnol. 40, 817–820.

23. Amberger, J.S., Bocchini, C.A., Schiettecatte, F., Scott, A.F., and Hamosh, A. (2015). OMIM.org: Online Mendelian Inheritance in Man (OMIM®), an online catalog of human genes and genetic disorders. Nucleic Acids Res. 43, D789–D798.

24. Danis, D., Bamshad, M.J., Bridges, Y., Cacheiro, P., Carmody, L.C., Chong, J.X., Coleman, B., Dalgleish, R., Freeman, P.J., Graefe, A.S.L., et al. (2024). A corpus of GA4GH Phenopackets: case-level phenotyping for genomic diagnostics and discovery.

25. Harada, Y., Sakamoto, T., Sugimoto, S., and Shimizu, T. (2024). Longitudinal Changes in Diagnostic Accuracy of a Differential Diagnosis List Developed by an AI-Based Symptom Checker: Retrospective Observational Study. JMIR Form Res 8, e53985.

26. Liu, X., Liu, H., Yang, G., Jiang, Z., Cui, S., Zhang, Z., Wang, H., Tao, L., Sun, Y., Song, Z., et al. (2025). A generalist medical language model for disease diagnosis assistance. Nat. Med. 31, 932– 942.

27. 100,000 Genomes Project Pilot Investigators, Smedley, D., Smith, K.R., Martin, A., Thomas, E.A., McDonagh, E.M., Cipriani, V., Ellingford, J.M., Arno, G., Tucci, A., et al. (2021). 100,000 Genomes Pilot on Rare-Disease Diagnosis in Health Care - Preliminary Report. N. Engl. J. Med. 385, 1868– 1880.

28. Shefchek, K.A., Harris, N.L., Gargano, M., Matentzoglu, N., Unni, D., Brush, M., Keith, D., Conlin, T., Vasilevsky, N., Zhang, X.A., et al. (2020). The Monarch Initiative in 2019: an integrative data and analytic platform connecting phenotypes to genotypes across species. Nucleic Acids Res. 48, D704– D715.

29. Bridges, Y.S., de Souza, V., Cortes, K.G., Haendel, M., Harris, N.L., Korn, D.R., Marinakis, N.M., Matentzoglu, N., McLaughlin, J.A., Mungall, C.J., et al. (2024). Towards a standard benchmark for variant and gene prioritisation algorithms: PhEval - Phenotypic inference Evaluation framework.

30. Temsah, M.-H., Jamal, A., Alhasan, K., Temsah, A.A., and Malki, K.H. (2024). OpenAI o1-Preview vs. ChatGPT in healthcare: A new frontier in medical AI reasoning. Cureus 16, e70640.

31. Shieh, A., Tran, B., He, G., Kumar, M., Freed, J.A., and Majety, P. (2024). Assessing ChatGPT 4.0’s test performance and clinical diagnostic accuracy on USMLE STEP 2 CK and clinical case reports. Sci. Rep. 14, 9330.

32. Gemini Team, Anil, R., Borgeaud, S., Alayrac, J.-B., Yu, J., Soricut, R., Schalkwyk, J., Dai, A.M., Hauth, A., Millican, K., et al. (2023). Gemini: A family of highly capable multimodal models.

33. Chen, Z., Cano, A.H., Romanou, A., Bonnet, A., Matoba, K., Salvi, F., Pagliardini, M., Fan, S., Köpf, A., Mohtashami, A., et al. (2023). MEDITRON-70B: Scaling medical pretraining for large language models.

34. Jacobsen, J.O.B., Kelly, C., Cipriani, V., Research Consortium, G.E., Mungall, C.J., Reese, J., Danis, D., Robinson, P.N., and Smedley, D. (2022). Phenotype-driven approaches to enhance variant prioritization and diagnosis of rare disease. Hum. Mutat. 43, 1071–1081.

35. Hager, P., Jungmann, F., Holland, R., Bhagat, K., Hubrecht, I., Knauer, M., Vielhauer, J., Makowski, M., Braren, R., Kaissis, G., et al. (2024). Evaluation and mitigation of the limitations of large language models in clinical decision-making. Nat. Med.

36. Klement, W., and El Emam, K. (2023). Consolidated Reporting Guidelines for Prognostic and Diagnostic Machine Learning Modeling Studies: Development and Validation. J. Med. Internet Res. 25, e48763.

