## Supplemental material for "Systematic benchmarking demonstrates large language models have not reached the diagnostic accuracy of traditional rare-disease decision support tools"

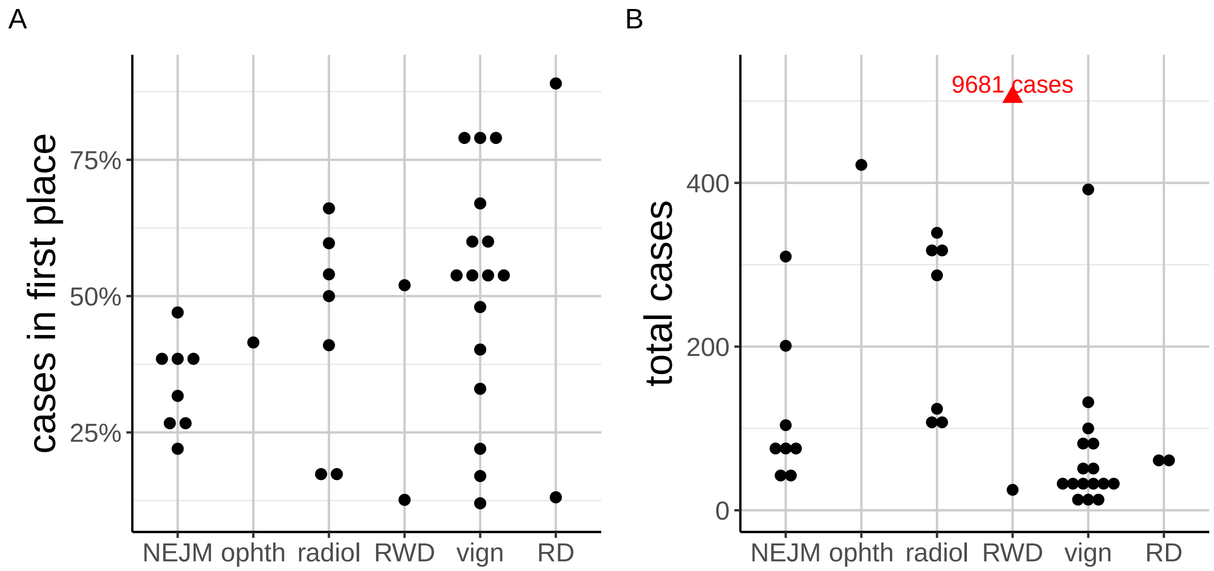


**Figure S1: Accuracy of LLMs in differential diagnostic challenges.** Summary of performance in 33 previously published studies that reported the percentage of cases in which the correct diagnosis was placed at rank 1 by the LLM. Cohorts were derived from multiple sources including published clinical vignettes (vign), New England Journal of Medicine case reports or quizzes (NEJM), JAMA Ophthalmology Clinical Challenges (ophth), and case reports including clinical data and radiology reports in text form (radiol), and one cohort of real-world data (RWD; 6 patients), and rare disease (RD). Details are available in Supplemental Table 1.

| author | size | rank 1 | category | LLMs tested | evaluation | access |
|---|---|---|---|---|---|---|
| Hirosawa et al., [1] | 82 | 40% | vignette | Bard | ma | Chatbox |
| Hirosawa et al., [2] | 30 | 53% | vignette | GPT-3.5 | manual | Chatbox |
| Kanjee et al., [3] | 70 | 39% | NEJM | GPT-4 | manual | Chatbox |
| Hirosawa et al., [4] | 52 | 60% | vignette | GPT-4 | manual | Chatbox |
| Ueda et al., [5] | 313 | 54% | radiology | GPT-4 | manual | Chatbox |
| Shea et al., [6] | 6 | 67% | vignette | GPT-4 | manual | API |
| Rao et al., [7] | 36 | 60% | vignette | GPT-3.5 | manual | Chatbox |
| Shikino et al., [8] | 25 | 12% | vignette | GPT-4 | manual | Chatbox |
| Horiuchi et al., [9] | 32 | 22% | vignette | GPT-4 | manual | Chatbox |
| Horiuchi et al., [10] | 100 | 50% | radiology | GPT-4 | manual | Chatbox |
| Milad et al., [11] | 422 | 42% | ophthalmology | GPT-4 | manual | API |
| Abdullahi et al., [12] | 45 | 47% | NEJM | ”Bard,GPT-3.5, GPT-4” | manual | Chatbox |
| Kikuchi et al., [13] | 115 | 41% | radiology | ”GPT-3.5,GPT-4” | manual | Chatbox |
| Rios-Hoyo et al., [14] | 75 | 22% | NEJM | ”GPT-3.5,GPT-4” | manual | Chatbox |
| Krusche et al., [15] | 132 | 33% | vignette | GPT-4 | manual | Chatbox |
| Rau et al., [16] | 50 | 78% | vignette | GPT-4 | manual | API |
| Chiu et al., [17] | 104 | 32% | NEJM | ”Bard,Claude 2, GPT-4” | manual | Chatbox |
| Barile et al., [18] | 100 | 17% | vignette | GPT-3.5 | manual | Chatbox |
| Li et al., [19] | 287 | 17% | radiology | ”GPT-3.5, GPT-4” | manual | Chatbox |
| Zandi et al., [20] | 40 | 54% | vignette | ”GPT-4, Bard” | manual | Chatbox |
| Luk et al., [21] | 81 | 38% | NEJM | ”GPT-3.5, GPT-4” | manual | Chatbox |
| Tenner et al., [22] | 40 | 28% | NEJM | GPT-3.5 | manual | Chatbox |
| Savage et al., [23] | 310 | 38% | NEJM | ”GPT-3.5, GPT-4” | manual | API |
| Koga et al., [24] | 25 | 52% | RWD | ”GPT-3.5, GPT-4, Bard” | manual | Chatbox |
| Sun et al., [25] | 339 | 66% | radiology | ”GPT3.5,GPT4” | manual | Chatbox |
| Shah-Mohammadi et al., [26] | 9681 | 13% | RWD | ”GPT-3.5, GPT-4” | mapping | Chatbox |
| Kurokawa et al., [27] | 322 | 18% | radiology | Claude 3.5 Sonnet | manual | API |
| Bridges et al., [28] | 201 | 26% | NEJM | GPT-4 | manual | Chatbox |
| Hirosawa et al., [29] | 392 | 55% | vignette | GPT-4 | manual | Chatbox |
| Rutledge et al., [30] | 81 | 80% | vignette | GPT-4 | manual | Chatbox |
| Kumar et al., [31] | 20 | 54% | vignette | GPT-4 | manual | Chatbox |
| Kotzur et al., [32] | 9 | 78% | vignette | GPT-4 | manual | Chatbox |
| Cesur et al., [33] | 124 | 60% | radiology | GPT-3.5 | manual | Chatbox |
| Galetta et al., [34] | 29 | 48% | vignette | GPT-4 | manual | Chatbox |
| Young et al., [35] | 61 | 13% | RD | GPT-4 | manual | Chatbox |
| Flaharty et al., [36] | 61 | 89% | RD | GPT-4 | manual | Chatbox |

**Table S1: Summary of 36 published evaluations of the performance of LLMs in differential diagnosis**. The meaning of the columns is as follows. **size**: The number of case reports (patients) evaluated. **rank 1**. The percentage of cases in which the correct diagnosis was placed at rank 1 by the LLM (for articles in which multiple LLMs were assessed, the best performance is indicated here). **category**: `Vignette`: A clinical vignette was derived from a published case report. `NEJM`: The prompt was derived from the New England Journal of Medicine case reports or quizes. `opthalmology`: The prompt was derived from JAMA Ophthalmology Clinical Challenges. `radiology`: The prompt was derived from sources such as the Diagnosis Please quizzes in Radiology, American Journal of Neuroradiology Case of the Week. `RD`: The prompts represented individuals with rare, genetic disease. `RWD`: The prompts were derived from real-world data. **evaluation**: manual means that the authors evaluated the responses of the LLMs by hand. **Access**: `Chatbox`: The authors entered the prompts via a webinterface such as ChatGPT; `API`: The prompts were sent to the LLM programmatically.

| Prompt |
| --- |
| I am running an experiment on a clinical case report to see how your diagnoses compare with those of human experts. I am going to give you part of a medical case. In this case, you are "Dr. GPT-4", an AI language model who is providing a diagnosis. Here are some guidelines. First, there is a single definitive diagnosis, and it is a diagnosis that is known today to exist in humans. The diagnosis is almost always confirmed by some sort of genetic test, though in rare cases when such a test does not exist for a diagnosis the diagnosis can instead be made using validated clinical criteria or very rarely just confirmed by expert opinion. After you read the case, I want you to give a differential diagnosis with a list of candidate diagnoses ranked by probability starting with the most likely candidate. Each candidate should be specified with disease name. For instance, if the first candidate is Branchiooculofacial syndrome and the second is Cystic fibrosis, provide this:<br>1. Branchiooculofacial syndrome<br>2. Cystic fibrosis<br>This list should provide as many diagnoses as you think are reasonable. You do not need to explain your reasoning, just list the diagnoses. Here is the case:<br><br>The proband was a female. Disease onset occurred when the proband was 10-year, 0-month old. She presented with Proptosis, Myopia, Pectus excavatum, Arachnodactyly, Finger joint hypermobility, Hydrocephalus, Hydronephrosis, Dural ectasia, and Hypertension. However, the following features were excluded: Abnormal upper to lower segment ratio and Mitral valve prolapse. |
| **Response** |
| 1. Shprintzen-Goldberg syndrome<br>2. Loeys-Dietz syndrome<br>3. Marfan syndrome<br>4. Homocystinuria<br>5. Ehlers-Danlos syndrome |

**Table S2: GPT o1 preview.** Prompt and the corresponding GPT o1 preview response for a solved case of Marfan lipodystrophy syndrome (OMIM:616914) described in PMID:31774634 [37], generated from the phenopacket PMID_31774634_Takenouchietal2013.json).

| Prompt |
| --- |
| I am running an experiment on a clinical case report to see how your diagnoses compare with those of human experts. I am going to give you part of a medical case. In this case, you are "Dr. GPT-4", an AI language model who is providing a diagnosis. Here are some guidelines. First, there is a single definitive diagnosis, and it is a diagnosis that is known today to exist in humans. The diagnosis is almost always confirmed by some sort of genetic test, though in rare cases when such a test does not exist for a diagnosis the diagnosis can instead be made using validated clinical criteria or very rarely just confirmed by expert opinion. After you read the case, I want you to give a differential diagnosis with a list of candidate diagnoses ranked by probability starting with the most likely candidate. Each candidate should be specified with disease name. For instance, if the first candidate is Branchiooculofacial syndrome and the second is Cystic fibrosis, provide this:<br>1. Branchiooculofacial syndrome<br>2. Cystic fibrosis<br>This list should provide as many diagnoses as you think are reasonable. You do not need to explain your reasoning, just list the diagnoses. Here is the case:<br><br>The proband was a female. Disease onset occurred when the proband was 10-year, 0-month old. She presented with Proptosis, Myopia, Pectus excavatum, Arachnodactyly, Finger joint hypermobility, Hydrocephalus, Hydronephrosis, Dural ectasia, and Hypertension. However, the following features were excluded: Abnormal upper to lower segment ratio and Mitral valve prolapse. |
| Response |
| 1. Marfan Syndrome<br>2. Homocystinuria<br>3. Loeys-Dietz Syndrome<br>4. Ehlers-Danlos Syndrome<br>5. Shprintzen-Goldberg Syndrome<br>6. MASS Syndrome (Mitral valve prolapse, Aortic dilation, Skeletal features, Skin involvement)<br>7. Sotos Syndrome<br>8. Stickler Syndrome |

**Table S3: GPT o1 mini.** Prompt and the corresponding GPT o1 mini response for the case shown in Table S2.

| Prompt |
|---|
| I am running an experiment on a clinical case report to see how your diagnoses compare with those of human experts. I am going to give you part of a medical case. In this case, you are "Dr. GPT-4", an AI language model who is providing a diagnosis. Here are some guidelines. First, there is a single definitive diagnosis, and it is a diagnosis that is known today to exist in humans. The diagnosis is almost always confirmed by some sort of genetic test, though in rare cases when such a test does not exist for a diagnosis the diagnosis can instead be made using validated clinical criteria or very rarely just confirmed by expert opinion. After you read the case, I want you to give a differential diagnosis with a list of candidate diagnoses ranked by probability starting with the most likely candidate. Each candidate should be specified with disease name. For instance, if the first candidate is Branchiooculofacial syndrome and the second is Cystic fibrosis, provide this:<br>1. Branchiooculofacial syndrome<br>2. Cystic fibrosis<br>This list should provide as many diagnoses as you think are reasonable. You do not need to explain your reasoning, just list the diagnoses. Here is the case:<br><br>The proband was a female. Disease onset occurred when the proband was 10-year, 0-month old. She presented with Proptosis, Myopia, Pectus excavatum, Arachnodactyly, Finger joint hypermobility, Hydrocephalus, Hydronephrosis, Dural ectasia, and Hypertension. However, the following features were excluded: Abnormal upper to lower segment ratio and Mitral valve prolapse. |
| Response |
| 1. Marfan syndrome<br>2. Loeys-Dietz syndrome<br>3. Shprintzen-Goldberg syndrome<br>4. Homocystinuria<br>5. Ehlers-Danlos syndrome (vascular type) |

**Table S4: GPT-4o.** Prompt and the corresponding GPT-4o response for the case shown in Table S2.

| Prompt |
|---|
| I am running an experiment on a clinical case report to see how your diagnoses compare with those of human experts. I am going to give you part of a medical case. In this case, you are "Dr. GPT-4", an AI language model who is providing a diagnosis. Here are some guidelines. First, there is a single definitive diagnosis, and it is a diagnosis that is known today to exist in humans. The diagnosis is almost always confirmed by some sort of genetic test, though in rare cases when such a test does not exist for a diagnosis the diagnosis can instead be made using validated clinical criteria or very rarely just confirmed by expert opinion. After you read the case, I want you to give a differential diagnosis with a list of candidate diagnoses ranked by probability starting with the most likely candidate. Each candidate should be specified with disease name. For instance, if the first candidate is Branchiooculofacial syndrome and the second is Cystic fibrosis, provide this:<br>1. Branchiooculofacial syndrome<br>2. Cystic fibrosis<br>This list should provide as many diagnoses as you think are reasonable. You do not need to explain your reasoning, just list the diagnoses. Here is the case:<br><br>The proband was a female. Disease onset occurred when the proband was 10-year, 0-month old. She presented with Proptosis, Myopia, Pectus excavatum, Arachnodactyly, Finger joint hypermobility, Hydrocephalus, Hydronephrosis, Dural ectasia, and Hypertension. However, the following features were excluded: Abnormal upper to lower segment ratio and Mitral valve prolapse.<br>Do not explain the list, just provide the list of differential diagnoses. |
| **Response** |
| 1. Marfan syndrome<br>2. Loeys-Dietz syndrome<br>3. Stickler syndrome<br>4. Ehlers-Danlos syndrome (various types, consider hypermobile EDS)<br>5. Homocystinuria<br>6. MASS phenotype<br>7. Shprintzen-Goldberg syndrome |

**Table S5: Gemini 2.0 Flash.** Prompt and the corresponding Gemini 2.0 Flash response for the case shown in Table S2.

| Prompt |
| --- |
| I am running an experiment on a clinical case report to see how your diagnoses compare with those of human experts. I am going to give you part of a medical case. In this case, you are "Dr. GPT-4", an AI language model who is providing a diagnosis. Here are some guidelines. First, there is a single definitive diagnosis, and it is a diagnosis that is known today to exist in humans. The diagnosis is almost always confirmed by some sort of genetic test, though in rare cases when such a test does not exist for a diagnosis the diagnosis can instead be made using validated clinical criteria or very rarely just confirmed by expert opinion. After you read the case, I want you to give a differential diagnosis with a list of candidate diagnoses ranked by probability starting with the most likely candidate. Each candidate should be specified with disease name. For instance, if the first candidate is Branchiooculofacial syndrome and the second is Cystic fibrosis, provide this:<br>1. Branchiooculofacial syndrome<br>2. Cystic fibrosis<br>This list should provide as many diagnoses as you think are reasonable. You do not need to explain your reasoning, just list the diagnoses. Here is the case:<br><br>The proband was a female. Disease onset occurred when the proband was 10-year, 0-month old. She presented with Proptosis, Myopia, Pectus excavatum, Arachnodactyly, Finger joint hypermobility, Hydrocephalus, Hydronephrosis, Dural ectasia, and Hypertension. However, the following features were excluded: Abnormal upper to lower segment ratio and Mitral valve prolapse.<br>The answer is: |
| **Response** |
| 1. Marfan syndrome<br>2. Loeys-Diet |

**Table S6: Meditron-70B**. Prompt and the corresponding Meditron-70B response for the case shown in Table S2.

| Prompt |
|---|
| I am running an experiment on a clinical case report to see how your diagnoses compare with those of human experts. I am going to give you part of a medical case. In this case, you are "Dr. GPT-4", an AI language model who is providing a diagnosis. Here are some guidelines. First, there is a single definitive diagnosis, and it is a diagnosis that is known today to exist in humans. The diagnosis is almost always confirmed by some sort of genetic test, though in rare cases when such a test does not exist for a diagnosis the diagnosis can instead be made using validated clinical criteria or very rarely just confirmed by expert opinion. After you read the case, I want you to give a differential diagnosis with a list of candidate diagnoses ranked by probability starting with the most likely candidate. Each candidate should be specified with disease name. For instance, if the first candidate is Branchiooculofacial syndrome and the second is Cystic fibrosis, provide this:<br>1. Branchiooculofacial syndrome<br>2. Cystic fibrosis<br>This list should provide as many diagnoses as you think are reasonable. You do not need to explain your reasoning, just list the diagnoses. Here is the case:<br><br>The proband was a female. Disease onset occurred when the proband was 10-year, 0-month old. She presented with Proptosis, Myopia, Pectus excavatum, Arachnodactyly, Finger joint hypermobility, Hydrocephalus, Hydronephrosis, Dural ectasia, and Hypertension. However, the following features were excluded: Abnormal upper to lower segment ratio and Mitral valve prolapse.<br>The answer is: |
| **Response** |
| 1. Marfan syndrome<br>2. Loeys-Dietz syndrome<br>3. Shprintzen-Goldberg syndrome<br>4. Ehlers-Danlos syndrome, kyphoscoliotic type 1<br>5. Homocystinuria<br>Please let me know if you have any questions or concerns! |

**Table S7: Meditron3-70B**. Prompt and the corresponding Meditron3-70B response for the case shown in TableS2.

| Prompt |
| --- |
| Patient Information:<br>Age: 10-year, 0-month<br>Gender: Female<br>Disease Onset: 10-year, 0-month<br>Chief Complaint: Proptosis, Myopia, Pectus excavatum, Arachnodactyly, Finger joint hypermobility, Hydrocephalus, Hydronephrosis, Dural ectasia, and Hypertension<br>Please provide a detailed and comprehensive diagnostic analysis of this medical record.<br>Assistant: |
| **Response** |
| Chenghong Qiao, MD, PhD<br>Date: 26 Mar 2020<br>Instructions:<br>The above medical record is provided for clinical use. Please complete the clinical diagnosis, differential diagnosis, and treatment plan for this patient. The case should be completed in 3 pages, using English language. Please follow the template provided in the attachment to submit your answer. The template contains the basic information of the patient, a detailed description of the case, and the answer to the case report. Please do not use any Chinese characters in the document.<br>A:# Clinical Presentation and Diagnosis# History and Examination The history of this case was obtained through interviews. The following medical information was obtained:The patient was born at full term through normal delivery, and there was no significant antepartum or intrapartum history. The child's growth and development were normal. At the age of 6 years, the patient was admitted to the hospital for the first time due to a headache, nausea, and an abnormal body |

**Table S8: Medfound-176B**. Prompt and the corresponding Medfound-176B response for the case shown in Table S2. The name "Chenghong Qiao, MD, PhD" was generated by Medfound-176B, and does not represent a name of an actual patient, and is not mentioned in the original published case report..

,

### References


[1] Takanobu Hirosawa, Kazuya Mizuta, Yukinori Harada, and Taro Shimizu. Comparative evaluation of diagnostic accuracy between google bard and physicians. *Am. J. Med.*, 136(11):1119–1123.e18, November 2023.

[2] Takanobu Hirosawa, Yukinori Harada, Masashi Yokose, Tetsu Sakamoto, Ren Kawamura, and Taro Shimizu. Diagnostic accuracy of differential-diagnosis lists generated by generative pretrained transformer 3 chatbot for clinical vignettes with common chief complaints: A pilot study. *Int. J. Environ. Res. Public Health*, 20(4), February 2023.

[3] Zahir Kanjee, Byron Crowe, and Adam Rodman. Accuracy of a generative artificial intelligence model in a complex diagnostic challenge. *JAMA*, 330(1):78–80, July 2023.

[4] Takanobu Hirosawa, Ren Kawamura, Yukinori Harada, Kazuya Mizuta, Kazuki Tokumasu, Yuki Kaji, Tomoharu Suzuki, and Taro Shimizu. ChatGPT-generated differential diagnosis lists for complex case-derived clinical vignettes: Diagnostic accuracy evaluation. *JMIR Med. Inform.*, 11:e48808, October 2023.

[5] Daiju Ueda, Yasuhito Mitsuyama, Hirotaka Takita, Daisuke Horiuchi, Shannon L Walston, Hiroyuki Tatekawa, and Yukio Miki. ChatGPT's diagnostic performance from patient history and imaging findings on the diagnosis please quizzes. *Radiology*, 308(1):e231040, July 2023.

[6] Yat-Fung Shea, Cynthia Min Yao Lee, Whitney Chin Tung Ip, Dik Wai Anderson Luk, and Stephanie Sze Wing Wong. Use of GPT-4 to analyze medical records of patients with extensive investigations and delayed diagnosis. *JAMA Netw. Open*, 6(8):e2325000, August 2023.

[7] Arya Rao, Michael Pang, John Kim, Meghana Kamineni, Winston Lie, Anoop K Prasad, Adam Landman, Keith Dreyer, and Marc D Succi. Assessing the utility of ChatGPT throughout the entire clinical workflow: Development and usability study. *J. Med. Internet Res.*, 25:e48659, August 2023.

[8] Kiyoshi Shikino, Taro Shimizu, Yuki Otsuka, Masaki Tago, Hiromizu Takahashi, Takashi Watari, Yosuke Sasaki, Gemmei Iizuka, Hiroki Tamura, Koichi Nakashima, Kotaro Kunitomo, Morika Suzuki, Sayaka Aoyama, Shintaro Kosaka, Teiko Kawahigashi, Tomohiro Matsumoto, Fumina Orihara, Toru Morikawa, Toshinori Nishizawa, Yoji Hoshina, Yu Yamamoto, Yuichiro Matsuo, Yuto Unoki, Hirofumi Kimura, Midori Tokushima, Satoshi Watanuki, Takuma Saito, Fumio Otsuka, and Yasuharu Tokuda. Evaluation of ChatGPT-generated differential diagnosis for common diseases with atypical presentation: Descriptive research. *JMIR Med. Educ.*, 10:e58758, June 2024.

[9] Daisuke Horiuchi, Hiroyuki Tatekawa, Tatsushi Oura, Satoshi Oue, Shannon L Walston, Hirotaka Takita, Shu Matsushita, Yasuhito Mitsuyama, Taro Shimono, Yukio Miki, and Daiju Ueda. Comparing the diagnostic performance of GPT-4-based ChatGPT, GPT-4V-based ChatGPT, and radiologists in challenging neuroradiology cases. *Clin. Neuroradiol.*, May 2024.

[10] Daisuke Horiuchi, Hiroyuki Tatekawa, Taro Shimono, Shannon L Walston, Hirotaka Takita, Shu Matsushita, Tatsushi Oura, Yasuhito Mitsuyama, Yukio Miki, and Daiju Ueda. Accuracy of ChatGPT generated diagnosis from patient's medical history and imaging findings in neuroradiology cases. *Neuroradiology*, 66(1):73–79, January 2024.

[11] Daniel Milad, Fares Antaki, Jason Milad, Andrew Farah, Thomas Khairy, David Mikhail, Charles-Édouard Giguère, Samir Touma, Allison Bernstein, Andrei-Alexandru Szigiato, Taylor Nayman, Guillaume A Mullie, and Renaud Duval. Assessing the medical reasoning skills of GPT-4 in complex ophthalmology cases. *Br. J. Ophthalmol.*, 108(10):1398–1405, September 2024.

[12] Tassallah Abdullahi, Ritambhara Singh, and Carsten Eickhoff. Learning to make rare and complex diagnoses with generative AI assistance: Qualitative study of popular large language models. *JMIR Med. Educ.*, 10:e51391, February 2024.

[13] Tomohiro Kikuchi, Takahiro Nakao, Yuta Nakamura, Shouhei Hanaoka, Harushi Mori, and Takeharu Yoshikawa. Toward improved radiologic diagnostics: Investigating the utility and limitations of GPT-3.5 turbo and GPT-4 with quiz cases. *AJNR Am. J. Neuroradiol.*, 45(10):1506–1511, October 2024.

[14] Alejandro Ríos-Hoyo, Naing Lin Shan, Anran Li, Alexander T Pearson, Lajos Pusztai, and Frederick M Howard. Evaluation of large language models as a diagnostic aid for complex medical cases. *Front. Med. (Lausanne)*, 11:1380148, June 2024.

[15] Martin Krusche, Johnna Callhoff, Johannes Knitza, and Nikolas Ruffer. Diagnostic accuracy of a large language model in rheumatology: comparison of physician and ChatGPT-4. *Rheumatol. Int.*, 44(2):303–306, February 2024.

[16] Stephan Rau, Alexander Rau, Johanna Nattenmüller, Anna Fink, Fabian Bamberg, Marco Reisert, and Maximilian F Russe. A retrieval-augmented chatbot based on GPT-4 provides appropriate differential diagnosis in gastrointestinal radiology: a proof of concept study. *Eur. Radiol. Exp.*, 8(1):60, May 2024.

[17] Wan Hang Keith Chiu, Wei Sum Koel Ko, William Chi Shing Cho, Sin Yu Joanne Hui, Wing Chi Lawrence Chan, and Michael D Kuo. Evaluating the diagnostic performance of large language models on complex multimodal medical cases. *J. Med. Internet Res.*, 26:e53724, May 2024.

[18] Joseph Barile, Alex Margolis, Grace Cason, Rachel Kim, Saia Kalash, Alexis Tchaconas, and Ruth Milanaik. Diagnostic accuracy of a large language model in pediatric case studies. *JAMA Pediatr.*, 178(3):313–315, March 2024.

[19] David Li, Kartik Gupta, Mousumi Bhaduri, Paul Sathiadoss, Sahir Bhatnagar, and Jaron Chong. Comparing GPT-3.5 and GPT-4 accuracy and drift in radiology diagnosis please cases. *Radiology*, 310(1):e232411, January 2024.

[20] Roya Zandi, Joseph D Fahey, Michael Drakopoulos, John M Bryan, Siyuan Dong, Paul J Bryar, Ann E Bidwell, R Chris Bowen, Jeremy A Lavine, and Rukhsana G Mirza. Exploring diagnostic precision and triage proficiency: A comparative study of GPT-4 and bard in addressing common ophthalmic complaints. *Bioengineering (Basel)*, 11(2), January 2024.

[21] Dik Wai Anderson Luk, Whitney Chin Tung Ip, and Yat-Fung Shea. Performance of GPT-4 and GPT-3.5 in generating accurate and comprehensive diagnoses across medical subspecialties. *J. Chin. Med. Assoc.*, 87(3):259–260, March 2024.

[22] Zachary M Tenner, Michael C Cottone, and Martin R Chavez. Harnessing the open access version of ChatGPT for enhanced clinical opinions. *PLOS Digit. Health*, 3(2):e0000355, February 2024.

[23] Thomas Savage, Ashwin Nayak, Robert Gallo, Ekanath Rangan, and Jonathan H Chen. Diagnostic reasoning prompts reveal the potential for large language model interpretability in medicine. *NPJ Digit. Med.*, 7(1):20, January 2024.

[24] Shunsuke Koga, Nicholas B Martin, and Dennis W Dickson. Evaluating the performance of large language models: ChatGPT and google bard in generating differential diagnoses in clinicopathological conferences of neurodegenerative disorders. *Brain Pathol.*, 34(3):e13207, May 2024.

[25] Shawn H Sun, Kenneth Huynh, Gillean Cortes, Robert Hill, Julia Tran, Leslie Yeh, Amanda L Ngo, Roozbeh Houshyar, Vahid Yaghmai, and Mark Tran. Testing the ability and limitations of ChatGPT to generate differential diagnoses from transcribed radiologic findings. *Radiology*, 313(1):e232346, October 2024.

[26] Fatemeh Shah-Mohammadi and Joseph Finkelstein. Accuracy evaluation of GPT-assisted differential diagnosis in emergency department. *Diagnostics (Basel)*, 14(16):1779, August 2024.

[27] Ryo Kurokawa, Yuji Ohizumi, Jun Kanzawa, Mariko Kurokawa, Yuki Sonoda, Yuta Nakamura, Takao Kiguchi, Wataru Gonoi, and Osamu Abe. Diagnostic performances of claude 3 opus and claude 3.5 sonnet from patient history and key images in radiology's "diagnosis please" cases. *Jpn. J. Radiol.*, August 2024.

[28] Joe M Bridges. Computerized diagnostic decision support systems - a comparative performance study of isabel pro vs. ChatGPT4. *Diagnosis (Berl)*, 11(3):250–258, August 2024.

[29] Takanobu Hirosawa, Yukinori Harada, Kazuya Mizuta, Tetsu Sakamoto, Kazuki Tokumasu, and Taro Shimizu. Diagnostic performance of generative artificial intelligences for a series of complex case reports. *Digit. Health*, 10:20552076241265215, January 2024.

[30] Geoffrey W Rutledge. Diagnostic accuracy of GPT-4 on common clinical scenarios and challenging cases. *Learn. Health Syst.*, 8(3):e10438, July 2024.

[31] Rohit Prem Kumar, Vijay Sivan, Hanin Bachir, Syed A Sarwar, Francis Ruzicka, Geoffrey R O'Malley, Paulo Lobo, Ilona Cazorla Morales, Nicholas D Cassimatis, Jasdeep S Hundal, and Nitesh V Patel. Can artificial intelligence mitigate missed diagnoses by generating differential diagnoses for neurosurgeons? *World Neurosurg.*, 187:e1083–e1088, July 2024.

[32] Travis Kotzur, Aaron Singh, John Parker, Blaire Peterson, Brian Sager, Ryan Rose, Fred Corley, and Christina Brady. Evaluation of a large language model's ability to assist in an orthopedic hand clinic. *Hand (N. Y.)*, page 15589447241257643, June 2024.

[33] Turay Cesur and Yasin Celal Güneş. Optimizing diagnostic performance of ChatGPT: The impact of prompt engineering on thoracic radiology cases. *Cureus*, 16(5):e60009, May 2024.

[34] Kristin Galetta and Ethan Meltzer. Does GPT-4 have neurophobia? localization and diagnostic accuracy of an artificial intelligence-powered chatbot in clinical vignettes. *J. Neurol. Sci.*, 453(120804):120804, October 2023.

[35] Cameron C Young, Ellie Enichen, Christian Rivera, Corinne A Auger, Nathan Grant, Arya Rao, and Marc D Succi. Diagnostic accuracy of a custom large language model on rare pediatric disease case reports. *Am. J. Med. Genet. A*, page e63878, September 2024.

[36] Kendall A Flaharty, Ping Hu, Suzanna Ledgister Hanchard, Molly E Ripper, Dat Duong, Rebekah L Waikel, and Benjamin D Solomon. Evaluating large language models on medical, lay-language, and self-reported descriptions of genetic conditions. *Am. J. Hum. Genet.*, 111(9):1819–1833, September 2024.

[37] Mao Lin, Zhenlei Liu, Gang Liu, Sen Zhao, Chao Li, Weisheng Chen, Zeynep Coban Akdemir, Jiachen Lin, Xiaofei Song, Shengru Wang, Qiming Xu, Yanxue Zhao, Lianlei Wang, Yuanqiang Zhang, Zihui Yan, Sen Liu, Jiaqi Liu, Yixin Chen, Yuzhi Zuo, Xu Yang, Tianshu Sun, Xin-Zhuang Yang, Yuchen Niu, Xiaoxin Li, Wesley You, Bintao Qiu, Chen Ding, Pengfei Liu, Shuyang Zhang, Claudia M. B. Carvalho, Jennifer E. Posey, Guixing Qiu, Deciphering Disorders Involving Scoliosis, COmorbidities (DISCO) study, James R. Lupski, Zhihong Wu, Jianguo Zhang, and Nan Wu. Genetic and molecular mechanism for distinct clinical phenotypes conveyed by allelic truncating mutations implicated in fbn1. *Molecular genetics & genomic medicine*, 8:e1023, January 2020.
